# Causal association of long COVID with brain structure changes: Findings from a 2-sample Mendelian randomization study

**DOI:** 10.1101/2025.02.12.25322170

**Authors:** Hui Li, Yihe Yang, Pingjian Ding, Rong Xu

**Affiliations:** Center For Artificial Intelligence in Drug Discovery, Sears Tower T304, School of Medicine, Case Western Reserve University, 10900 Euclid Avenue, Cleveland, OH 44106, United States of America; Department of Population and Quantitative Health Sciences, Case Western Reserve University, Cleveland, Ohio, USA

**Keywords:** brain structure, causal association, *cis*-Mendelian Randomization, long COVID

## Abstract

Nearly 7.5% U.S. adults have long COVID. Recent epidemiological studies indicated that long COVID, is significantly associated with subsequent brain structure changes. However, it remains unknown if long COVID is causally associated with brain structure change. Here we applied two Mendelian Randomization (MR) methods – Inverse Variance Weighting MR method (IVW) for correlated instrument variables and Component analysis-based Generalized Method of Moments (PC-GMM) – to examine the potential causal relationships from long COVID to brain structure changes. The MR study was based on an instrumental variable analysis of data from a recent long COVID genome-wide association study (GWAS) (3,018 cases and 994,582 controls), the Enhancing NeuroImaging Genetics through Meta Analysis (ENIGMA) (Global and regional cortical measures, N = 33,709; combined hemispheric subcortical volumes, N = 38,851), and UK Biobank (left/right subcortical volumes, N = 19,629). We found no significant causal relationship between long COVID and brain structure changes. As we gain more insights into long COVID and its long-term health outcomes, future works are necessary to validate our findings and understand the mechanisms underlying the observed associations, though not causal, of long COVID with subsequent brain structure changes.

## 1. Introduction

According to data from National Center for Health Statistics in 2022, about 7.5% of U.S. adults were living with long COVID, also known as post COVID-19 condition or postacute sequelae of COVID-19 (PASC) [1]. Long COVID is currently defined by World Health Organization as a range of symptoms that present after COVID-19 and persist after 3 months [2, 3]. Neuropsychiatric symptoms including depression, anxiety and cognitive deficits have been widely reported as symptoms in patients with long COVID across the severity spectrum of this respiratory disease [3, 4, 5]. Direct viral infection of the central nervous system [6, 7], or prolonged neuroinflammation [8] have been proposed as potential underlying mechanisms. Recent studies suggest that brain structure abnormalities are involved in long covid associated neuropsychiatric symptoms. Heine et al. [9] carried out a prospective cross-sectional observational study and showed that the volumes of the left thalamus, putamen and pallidum for long COVID with fatigue decreased. Michael et al. [3] implemented a prospective, national longitudinal study to investigate the pathophysiology and recovery trajectory of persistent long COVID cognitive deficits and found reduced anterior cingulate cortex volume one year after admission. These studies showed that long COVID was associated with subsequent brain structure changes. However, due to the inherent challenges related to confounding variables and reverse causation in epidemiological observational studies, it remains unknown whether long COVID leads to brain structure changes.

Mendelian randomization (MR) is a useful tool to investigate causal relationship. It utilizes genetic variants as instrument variables to randomly allocate an exposure and estimate the causal relationship of the exposure on an outcome. Recently Ding and Xu [10] carried out a large-scale MR study and showed that COVID-19 infection and severity were not causally associated with brain structure. Meanwhile, Zhou et al. [11] showed nominally significant causal effect of COVID-19 infection, hospitalized COVID-19 and severe COVID-19 on specific brain structures. Currently, it remains unknown if long COVID is causally associated with subsequent brain structure changes. Genetic susceptibility of long COVID is different from COVID. Specifically, the genome-wide significant SNPs in long COVID [2] are not genome-wide significant in GWAS for COVID infection (N=1,348,701) and COVID severity (N=1,557,411)(https://www.covid19hg.org/results/r5/) [10]. Most COVID patients recover within 3 months and only a subset of them continue to have long COVID, approximately 14% to 43% [12, 13, 14]. Therefore, in this study we aim to investigate the causal relationship of long COVID with brain structure change including both cortical and subcortical brain structures.

## 2. Method

### 2.1 Datasets

In this study, we used GWAS summary statistics of long COVID [2] (N = 3,018 for cases and 994,582 for controls, available from https://www.medrxiv.org/node/671137. external-links.html) and GWAS summary statistics of brain structure (global and regional cortices [15], N = 33,709, available from http://enigma.ini.usc.edu/research/download-enigma-gwas-results; combined hemispheric subcortical regions [16], N = 38,851, available from http://enigma.ini.usc.edu/research/download-enigma-gwas-r and left/right subcortices [17], N = 19,629, available from https://github.com/BIG-S2/GWAS). More details are in Supplementary Table 1.

For long COVID GWAS, we used the strict broad case control meta analysis between long COVID patients with an earlier test-verified severe acute respiratory syndrome coronavirus 2 (SARS-CoV-2) infection (i.e. strict cases) and genetically ancestry-matched population controls without known long COVID (i.e. broad controls). Long COVID was defined as patients with any symptoms that present after COVID-19 infection and persist after three months. The most common symptoms were fatigue, anosmia, shortness of breath, persistent cough and problems with memory and concentration.

The GWAS investigations of brain structures were conducted with participants in ENIGMA and UK Biobank. The outcome measures include surface area and thickness of the global and 34 regional cortices [15] as well as volumes of 7 combined hemispheric [16] and 17 left/right subcortices [17], which are specifically amygdala, hippocampus, accumbens, putamen, pallidum, thalamus, insula, caudate, and brain stem. Included GWAS studies have obtained ethical approval from the corresponding ethics review boards.

### 2.2 Two sample *cis*-MR

Two *cis*-MR methods were adopted – IVW for correlated instrument variables [18] and PCGMM [19]. In total 16 Instrument variables within 3Mb around the identified lead variant rs9367106 in the long COVID GWAS study [2] were selected with p-value threshold of 5 × 10^−6^. To avoid numerically unstable causal effect estimate in the IVW method [20], LD clumping [21] was used to select informative SNPs and remove other correlated SNPs with *r*^2^ > 0.8 within a window size of 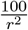 where *r*^2^ = 0.8. The window size was set based on the recommendation of the snp clumping in the “bigsnpr” package [22]. For both IVW and PCGMM, we searched appropriate proxies for those instrument SNPs not available in the brain structure outcome datasets with *r*^2^ = 0.6; we also harmonized long COVID and brain structure data and removed palindromic SNPs with minor allele frequency above 0.42 (see Supplementary Tables 2-4 for proxy SNPs and harmonized GWAS summary statistics for exposure and outcome.). For ivw, 3 informative SNPs were selected. According to PheWeb (https://pheweb.org/UKB-SAIGE/about) [23] and GWAS Catalog [24], none of the 3 instrument variables had pleiotropy effects through other risk factors. The mr_ivw function from MendelianRandomization package [18] was applied for ivw analysis. For PCGMM, we utilized the “mr_pcgmm” function from the MendelianRandomization package. As recommended by the function, LD clumping was carried out with parameter *r*^2^ > 0.95 within a window size of 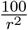 where *r*^2^ = 0.95. In total 4 SNPs were selected and based on PheWeb and GWAS Catalog, none of them were genome-wide significantly associated with pleiotropy risk factors. The selected 4 instrument SNPs were linearly combined and the first 2 principle components which explained 99% variance of the genetic data were chosen. For both ivw and pcgmm, F statistic was larger than 10, suggesting no weak instruments (see Supplementary Table 5) [25, 26].

### 2.3 MR multiple testing correction procedures

We accounted for multiple testing by defining significance at a false discovery rate (FDR) of 0.05. We performed separate FDR corrections for each subgroup of brain structure MR analyses [27] using Benjamini–Hochberg procedure [28]. Specially, the subgroups are: 1) long COVID on global cortical brain structures, 2) long COVID on regional cortical brain structures, 3) long COVID on hemispheric combined subcortical brain structures, 4) long COVID on left/right subcortical brain structures. The corrected Pvalue = original Pvalue 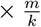, where m is the number of MR analyses in each subgroup and k is the rank of pvalue in a ascending order in that subgroup.

## 3. Results

Among the 94 brain structure summary statistics datasets (Sec. 2.1), the 7 combined subcortical volume datasets were excluded because only one instrument variable exists in the outcome datasets, even after proxy searching (Sec. 2.2). For the remaining 87 outcome datasets, we found no significant causal relationship between long COVID with brain structure changes in global cortex (Table 1), regional cortex (Figure 1B) and left/right subcortex (Table 1) under the control of FDR<0.05 for each sugroup (see Section MR multiple testing correction procedures ), i.e. the corrected Pvalue after FDR correction is larger than the significance threshold 0.05 for the surface area/thickness/volume of each specific brain region.

**Figure 1.**
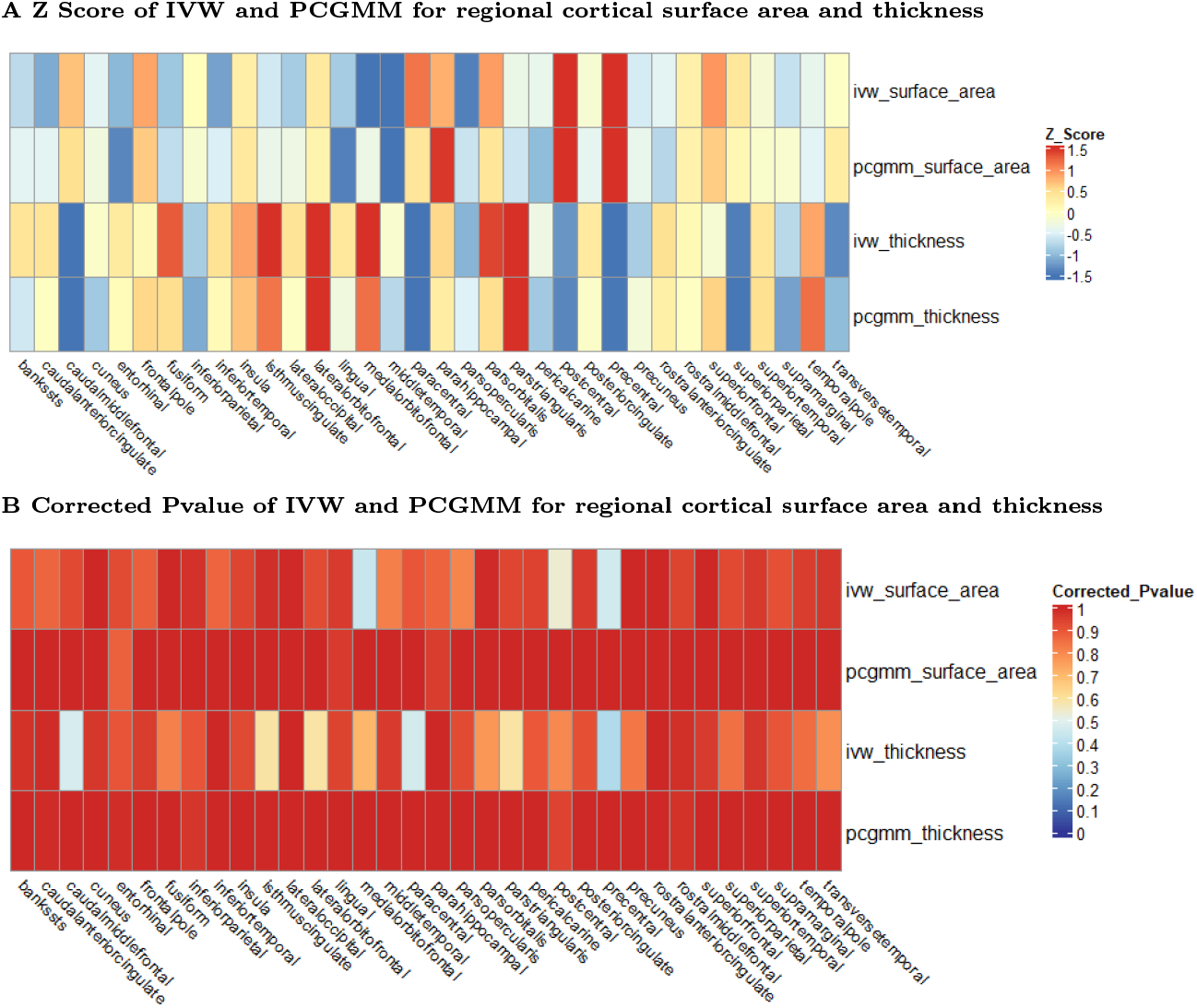
MR analysis for causal relationship of long COVID on regional cortical brain structure changes. Panel A shows the Z score of the IVW and PCGMM analysis for the causal relationship between long COVID and regional cortices, for both surface area and thickness. Panel B shows the correponding corrected Pvalue (Section MR multiple testing correction procedures ). Generally speaking, IVW and PCGMM produces similar Z scores and corrected P values. Both methods show that long COVID is not causally associated with regional cortical brain structure changes (i.e. corrected Pvalue > 0.05)

**Table 1.**
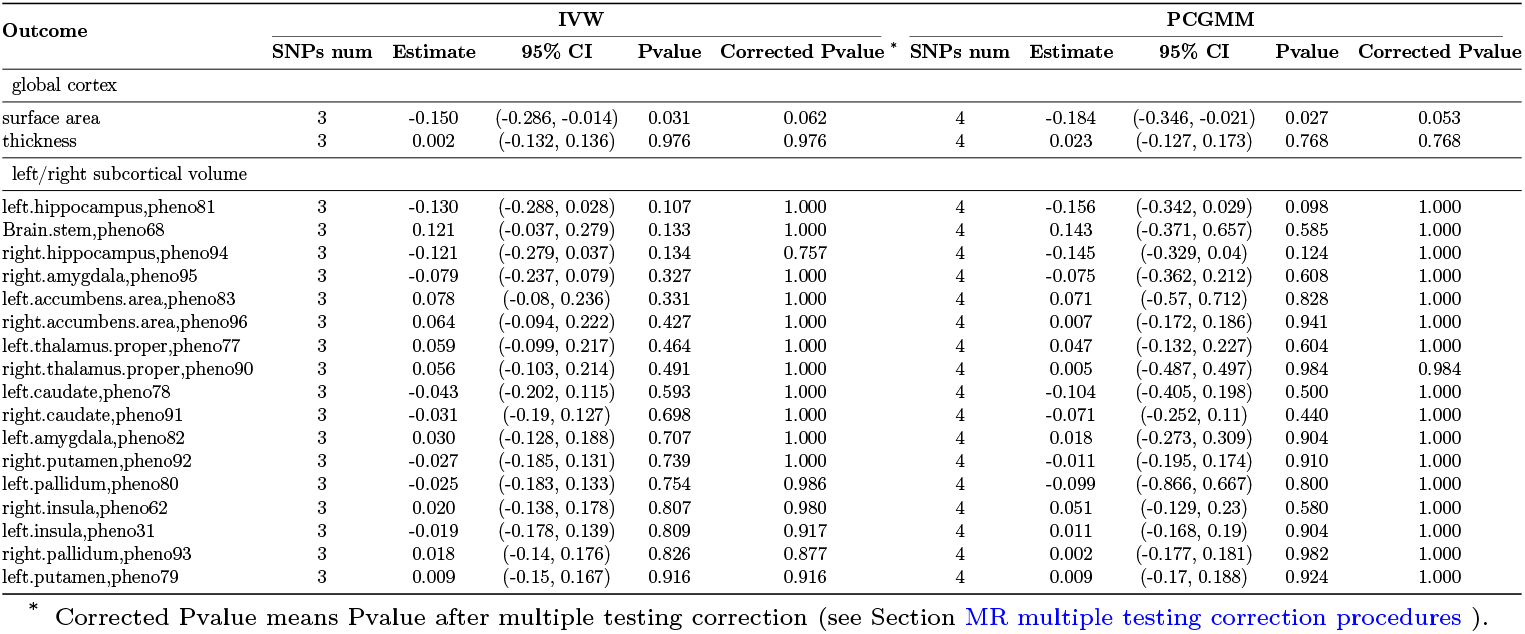
MR analysis for causal relationship of long COVID on global cortical and left/right subcortical brain structure changes. Both IVW and PCGMM show that long COVID is not causally associated with global cortical and left/right subcortical brain structure changes (i.e. corrected Pvalue *>* 0.05).

The null conclusion is consistent between IVW and PCGMM methods. Specifically, the sign of the causal effect estimates/z scores are mostly the same between IVW and PCGMM and the size of the causal effect estimates/z scores are largely similar (Table 1 for global cortices and left/right subcortices and Figure 1A for regional cortices), given that the two methods both work well.

## 4. Discussion

We implemented the first MR analysis to investigate the causal relationship between long COVID and brain structure changes, utilizing two cis-MR methods – IVW for correlated instruments and PCGMM. Both methods produced consistent results indicating that the genetic susceptibility of long COVID is not causally associated with brain structure changes.

Our MR analysis complements existing epidemiological observational studies and suggests that brain structure changes due to long COVID are dynamic and temporal. Recent epidemiological observational studies suggest long COVID is significantly associated with subsequent brain structure changes in specific brain regions. Heine et al. [9] showed decreased volumes of some subcortical brain regions for long COVID patients with fatigue. Michael et al. [3] found reduced volume for anterior cingulate cortex for long COVID patients one year after admission. However, these observational studies only report brain structure changes in a relatively short period after the acute SARS-CoV-2 infection, such as a median 7.5 months with inter-quartile range (first and third quartiles) 6.5–9.2 months [9] or one year after admission to hospital [3]. In contrast, MR estimates reflect a lifelong effect of genetic susceptibility of long COVID on brain structure changes. And there are epidemiological studies suggesting that some brain structure changes due to long COVID were discovered to recover. For example, Tian et al. [29] observed the dynamic brain changes from 3 to 10 months after discharge from hospital for 34 COVID patients and found that the cortical thickness were dynamic and finally returned to baseline. Du et al. [30] observed brain structure changes of COVID survivors at 1 year and 2 years after discharge from hospital and found that the decreased GMVs in the left middle frontal gyrus, inferior frontal gyrus of the operculum, right middle temporal gyrus, and inferior temporal gyrus returned to normal at two years.

Our findings about causal relationship between long COVID and brain structure changes is consistent with existing MR analyses between COVID and brain structure changes. Ding and Xu [10] showed that COVID-19 infection and severity were not causally associated with brain structure in a large-scale MR analysis. The GWAS samples for the exposure and outcome in this study are not overlapped. Bias due to weak instruments in a non-overlapping two-sample MR analysis is in the direction of the null and thus false positive findings can be avoided [31, 32]. Although Zhou et al. [11] proposed causal effect of COVID-19 infection, hospitalized COVID-19 and severe COVID-19 on specific brain structures, their results are only nominally significant.

Our study has limitations. First, there would be misclassification for a binary exposure (i.e. long COVID in this investigation) and collider bias “SARS-CoV-2 infection” may be present when analyzing long COVID cases with an earlier test-verified SARS-CoV-2 infection. Second, currently the only available GWAS study [2] for long COVID recruited patients with all symptoms rather than a specific symptom such as the neuropsychiatric symptom or fatigue. The selection criteria for symptoms matter because the brain structures are more likely to change for long COVID with neuropsychiatric symptom or fatigue than long COVID with symptoms less related to brain such as short of breath, or cough. Third, we did not examine the causal effects of long COVID on brain structures in subgroups stratified by age, gender, or comorbidities due to the lack of such GWAS data. Fourth, the GWAS summary statistics for both long COVID and brain structure changes in this study were primarily for individuals of European ancestry. The causal associations of long COVID and brain structure changes for ethnic and racial minorities in the US are important because they were disproportionately affected by the pandemic [33].

To validate our findings in the future, new large-scale GWAS studies for long COVID are necessary such as for a specific brain-related symptom of long COVID. GWAS studies of test-verified SARS-CoV-2 are needed for collider bias analysis if long COVID confirmation is conditioning on earlier test verification of COVID infection [34]. Epidemiological observational studies with much longer longitudinal follow-up periods are also recommended to investigate the long-term dynamic change of brain structure after COVID infection. Meanwhile, identifying the contributing factors for the temporal brain structure changes are also important since these factors are modifiable compared with genetic susceptibility and they could potentially be therapeutic targets to reverse the brain structure damage from long COVID earlier.

Taken together the complementary evidence from both epidemiological and MR studies, long-COVID is associated with subsequent brain structure changes. However this dynamic and temporal association relationship might be mediated by modifiable factors such as inflammation trigger by COVID infection [35, 36] other than genetic susceptibility. Targeting those modifiable mediators could potentially reverse the brain structure damage from long COVID. Future works are necessary to validate our findings and understand the underlying mechanisms of the association relationship between long COVID and subsequent brain structure changes.

## Author Contributions

R.X. conceptualized the study. H.L. , P.J.D. , Y.H.Y. designed the study. H.L. performed all experiments and wrote the manuscript. All authors critically contributed to the result interpretation and manuscript preparation. All authors read and approved the final manuscript.

## Ethics approval statement

This study relied on multiple publicly available GWAS summary statistics on long COVID and subcortical brain structure. Ethical approval was obtained in all original studies.

## Data availability

All the summary statistics in this study can be obtained from the original genome-wide association studies. Any other data generated during the analysis can be requested from the authors.

## Declaration of Competing Interest

The authors declare no conflicts of interest.

## Acknowledgments

The authors thank the participants from all cohorts who contributed to this study; and we are indebted to Dr. Xiaofeng Zhu for valuable discussions and suggestions. This work has been supported by NIH National Institute of Aging, USA R01 AG057557, R01 AG061388, R56 AG062272, National Institute on Alcohol Abuse and Alcoholism, USA (grant no. R01AA029831). This work was also partially supported by grant HG011052 and HG011052-03S1 from the National Human Genome Research Institute (NHGRI), USA.

## Notes

### Competing Interest Statement

The authors have declared no competing interest.

### Author Declarations

The source data were openly available before the initiation of the study. In this study, we used GWAS summary statistics of long COVID (Lammi et al., 2023) (N =3,018 for cases and 994,582 for controls, available from https://www.medrxiv.org/node/671137.external-links.html) and GWAS summary statistics of brain structure (global and regional cortices (Grasby et al., 2020), N = 33,709, available from http://enigma.ini.usc.edu/research/download-enigma-gwas-results; combined hemispheric subcortical regions (Satizabal et al., 2019), N = 38,851, available from http://enigma.ini.usc.edu/research/download-enigma-gwas-results; and left/right subcortices (Zhao et al., 2019), N = 19,629, available from https://github.com/BIG-S2/GWAS).

